# Monitoring influenza-like symptoms in the UK through participatory surveillance: insights from FluSurvey over two winter seasons (2023-24 and 2024-25)

**DOI:** 10.64898/2026.02.12.26345150

**Authors:** Rebecca E Green, Jonathon Mellor, Christopher Rawlinson, Elizabeth Waller, Nurin Abdul Aziz, Conall H Watson, Gavin Dabrera

## Abstract

FluSurvey is a participatory surveillance system used to monitor trends in influenza and other respiratory viruses through weekly symptom surveys among the UK population. We aimed to characterise the wider impact of “influenza-like illnesses” (ILI) among FluSurvey participants and assess correlations of ILI with other established influenza surveillance systems.

We included data reported by FluSurvey participants over the 2023-24 and 2024-25 winter seasons. Using weekly symptoms surveys, we derived ILI episodes and estimated the proportion reporting healthcare service use, medication use, impact on daily life, absenteeism and use of tests. We applied existing methodologies (omitting first report and weighting to the age-sex structure of England) and assessed cross-correlations of weekly FluSurvey ILI rates with the national surveillance of GP ILI consultations, influenza hospital admissions, and influenza PCR test positivity at time lags of up to +/− 2 weeks.

There were 3057 participants over two winter seasons (N_2023-24_=2540, 63% female, mean age 60 years; N_2024-25_=2273, 64% female, mean age 61 years). Of 1868 ILI episodes, only a minority contacted healthcare services (14%, most frequently visiting the GP). A large proportion of episodes reported medication use (89%), impact on daily life (75%) and missing school or work (47%). Notable differences in testing behaviour were apparent by season, with fewer reporting use of tests in 2024-25. FluSurvey ILI rates were strongly correlated with other influenza surveillance, predominantly leading GP ILI consultations (max r=0.73), coinciding with influenza hospital admissions (max r=0.88) and lagging influenza test positivity (max r=0.88).

The majority of ILI reported to FluSurvey do not contact healthcare due to symptoms but experienced wider impacts on daily life. FluSurvey ILI corresponds well with other national influenza surveillance and provides broader context on community illness, supplementing the monitoring of influenza activity for public health response.

## Introduction

Whilst seasonal influenza contributes to morbidity, mortality and subsequent pressures on healthcare services (1), infections vary in severity and most resolve in the community. Among milder infections, population-level impacts are still apparent through absenteeism and health-related quality of life (2). Understanding the spread and characteristics of infections in the population can inform epidemiological assessment and transmission dynamic modelling in seasonal and pandemic waves.

In the UK, influenza is monitored using numerous surveillance tools across the breadth of disease to inform timely public health response (3). Core surveillance systems use information from virological testing and healthcare attendance to monitor trends, providing critical insights on test-confirmed and moderate to severe cases. Participatory surveillance tools directly engage with communities to actively report data, and complement these by allowing an understanding of the wider burden of infections including non-medically attended cases (4). They may be able to detect trends before it is seen in other surveillance data, such as an increase in “influenza-like-illness” (ILI) – commonly used as a symptom-based proxy for influenza where testing is unavailable – and can provide important context on health-related behaviours including healthcare use (5). As participatory systems comprise near real-time crowdsourced data, they are low cost, timely, and flexible, although biases may arise through voluntary participation (5).

FluSurvey is a web-based participatory surveillance tool used to monitor trends in ILI and other respiratory illness through weekly symptom surveys (6). Set up in 2009 during the influenza A(H1N1) pandemic, it is the longest running UK symptoms survey and is used for influenza surveillance each winter. Based on a standardised format deployed across other countries in Europe, it is also able to facilitate harmonised international surveillance (7).

Although several FluSurvey studies have been conducted previously (8–13), no study has comprehensively profiled the cohort since the COVID-19 pandemic. This includes symptom reporting and health-related behaviours – which are largely unknown post-COVID-19 – as well as the interpretation of ILI in the context of influenza given shifts in circulating viruses (14,15). A detailed understanding of information captured by the system will allow insights into the burden of symptomatic illness in the community and facilitate the detection of unusual activity.

Here, we characterise symptom reporting, ILI burden and health-related behaviours among FluSurvey participants over winter seasons 2023-24 and 2024-25. We implement methods to address possible biases in the data and compare FluSurvey ILI rates with core influenza surveillance (general practitioner/family physician consultations, hospital admissions and PCR positivity) to understand its timeliness and validity in comparison to other established systems.

## Methods

### Participants

FluSurvey is an online surveillance system launched by the London School of Hygiene and Tropical Medicine in 2009 and is now managed by the UK Health Security Agency (UKHSA). In October 2023, FluSurvey relaunched on a new web platform (https://flusurvey.net/en).

Previous participants were invited to re-register and new sign-ups were encouraged through recruitment campaigns. We used data collected since this relaunch comprising the 2023-24 and 2024-25 seasons, which operated between the following dates:

- 2023-24 season: 25 weeks from week 44 (launched mid-week w/c 30 October 2023) to week 16 (w/c 15 April 2024)
- 2024-25 season: 20 weeks from week 47 (w/c 18 November 2024) to week 14 (w/c 31 March 2025)

Recruitment methods included email contact with registered participants, word of mouth, social media and blog posts. Registration is open year-round to anyone residing in the UK aged 18 years and over. Individuals can participate on behalf of household members including children.

### Data collection

On registration and at the start of each season, participants complete an “intake” survey. This can be updated at any point if responses change (e.g. following influenza vaccination) and includes questions on sociodemographics and risk factors for respiratory illness.

Participants may then complete their weekly “symptoms” survey which includes three questions asked to all users:

- Symptoms experienced in the past week (or since their last report), including:
  ○ None
  ○ Respiratory (cough, runny nose, sore throat, sneezing, shortness of breath, coloured sputum, chest pain, nose bleed),
  ○ Systemic (malaise, muscle/joint pain, chills, fever, loss of appetite, headache),
  ○ Gastrointestinal (nausea, stomach ache, diarrhoea, vomiting),
  ○ Ocular (sticky/itchy eyes [added in 2024-25], watery/bloodshot eyes),
  ○ Other (loss of smell or taste, other symptom)
- Self-reported health scores (0 to 100)
- Number of social contacts in the previous 24 hours* [*symptomatic subset only in 2023-24]

Where symptoms are reported, follow-up questions are included on whether symptoms were of sudden onset and symptom start/end dates. Questions on healthcare use, testing, medication, and impact on daily routine as a result of these symptoms are additionally asked (Text S1).

Participants are randomly assigned to a day on registration and reminded to complete the symptoms survey by email on the scheduled day each week. However, participation is voluntary and surveys may be submitted at any frequency.

### Data extraction

We extracted pseudonymised survey data from the FluSurvey webtool and deduplicated multiple responses per participant, retaining one intake survey per season and one symptoms survey in each participating week. In both instances we kept the most recent submission (12). Participants who submitted an intake survey and at least one symptoms survey were included in analysis.

### Data preparation

#### ILI/ARI case definition

For each symptoms survey, we derived ILI and Acute Respiratory Infection (ARI) according to the European Centre for Disease Prevention and Control (ECDC) case definitions (16).

ILI: A sudden onset of symptoms with at least one of four systemic symptoms (fever or feverishness, malaise, headache, joint/muscle pain) and at least one of three respiratory symptoms (cough, sore throat, shortness of breath).

ARI: A sudden onset of symptoms with at least one of the following respiratory symptoms (cough, sore throat, shortness of breath, runny/blocked nose, sneezing).

#### ILI episodes

Participants meeting the ILI case definition in consecutive reporting weeks were combined into one ILI episode. If no symptom survey was submitted in the previous week but the participant met the ILI case definition in their last report, these were combined where the symptom onset date was approximately similar (±3 days) and not more than three weeks had elapsed. Episodes were calculated among “active” participants who submitted two or more surveys in that season.

We collapsed symptoms survey responses during the illness episode into dichotomous variables defining never/ever reporting symptoms, healthcare use, medication, impact on daily routine and testing behaviour in each episode (see Text S1).

Symptoms included each of the 22 symptoms in the survey. We additionally grouped these into categories and assessed the proportion experiencing any gastrointestinal symptom, any ocular symptom, and loss of taste/smell to understand the prevalence of non-respiratory or systemic symptoms (see Data Collection).

Healthcare use included any and each of the following: National Health Service (NHS) 111 (a non-emergency telephone or online service) or use of other NHS online services, telephoning the GP practice (primary care), visiting the GP, visiting accident and emergency (A&E) or out of hours services, hospital admission, and telephoning/visiting another medical service. If visiting healthcare services were selected, the number of days between symptom onset and health service use was derived using the minimum time reported during the episode. We did not calculate this for rarer healthcare outcomes (reported in ≤5% of episodes in a season).

Medication included any and each of the following: painkillers, cough medication, antibiotics, homeopathy or other alternative medicine, COVID-19 therapeutics, influenza antivirals, and other medication.

One question regarding impact on daily routine was included in 2023-24, asking whether daily routine was modified with absence from work or school indicated. In 2024-25 this was updated to ask if daily routine was affected, and whether the participant missed work or school if they reported “yes”. Absenteeism was derived in both seasons among the subset in employment or education, which included those who reported daycare or education; self-employed; and part– or full-time employment in their profile questionnaire. Impact on daily routine was not derived for 2023-24 given the number of participants who selected “not applicable – I don’t go to work / school”.

Testing questions also differed by season. In 2023-24 participants were asked whether they tested for COVID-19 or influenza because of their symptoms, including the type of test used (Text S1). In 2024-25, this was updated to include one question reflecting use of tests and result (Text S1). We examined these separately, assessing use of SARS-CoV-2 and influenza tests in 2023-24, and use of any test and test result in 2024-25.

Finally, the change in health score during the illness was calculated as the percentage difference between the median health score where no symptoms were reported and the minimum health score reported during the illness episode (13).

#### Crude ILI and ARI prevalence rates

We summed the number of surveys meeting the ILI case definition each week using the date the survey was submitted. We then converted these to rates per 1000 FluSurvey participants based on the number of participants contributing a symptoms survey in that week. This indicator has been used for national surveillance to date (17).

We applied the same calculation to derive the crude weekly rates of ARI and symptom reporting per 1000 FluSurvey participants, using the metric of interest as the numerator (i.e. the number of weekly surveys meeting the ARI case definition reporting each symptom).

#### ILI indicator methodology

We derived two additional FluSurvey weekly ILI indicators, applying methods that aim to address different biases using near real-time data (18).

##### a) Crude ILI rates (+QC)

To minimise possible reporting bias introduced through participants’ propensity to register and/or report when unwell, we omitted the first week of participation from analyses, retaining only those who report more than once. Rates per 1000 were then derived using this analytical subset.

##### b) Weighted ILI rates (by age and sex)

To address over/under-representation by age and sex through self-selection, we adjusted rates to the age/sex structure of England. We calculated rates per 1000 by sex and age in eight strata, using the following age groups based on age at survey submission: under 45s; 45-54; 55-64; 65 and over in females and males. The wider “under 45s” age category was curated due to small numbers among the younger age groups and a relatively uncommon outcome (<10% at peak activity) (Table 1). Per-strata rates were multiplied by the proportion of each group represented among the population in England (19), and summed.

**Table 1.** FluSurvey participant characteristics in the 2023-24 and 2024-25 season. *National Census 2021 UK data* (23) *are presented alongside for comparisons*.

|  | 2023-2024 | 2024-2025 | Census |
| --- | --- | --- | --- |
|  | N = 2,540 | N = 2,273 |  |
| **Country - n (%)** |  |  |  |
| England | 2,237 (89%) | 1,999 (89%) | 84.4% |
| Wales | 80 (3.2%) | 87 (3.9%) | 4.6% |
| Scotland | 166 (6.6%) | 132 (5.9%) | 8.1% |
| Northern Ireland | 14 (0.6%) | 11 (0.5%) | 2.8% |
| Other | 6 (0%) | 6 (0%) | . |
| Missing | 37 | 38 |  |
| **Sex - n (%)** |  |  |  |
| Male | 932 (37%) | 801 (35%) | 48.9% |
| Female | 1,595 (63%) | 1,464 (64%) | 51.1% |
| Other | 13 (0.5%) | 8 (0.4%) | . |
| **Age (years) - mean (SD)** | 60 (15) | 61 (15) |  |
| **Age group - n (%)** |  |  |  |
| 0-17 | 50 (2.0%) | 32 (1.4%) | 20.7% |
| 18-24 | 22 (0.9%) | 19 (0.8%) | 8.3% |
| 25-34 | 67 (2.6%) | 61 (2.7%) | 13.4% |
| 35-44 | 217 (8.5%) | 209 (9.2%) | 12.9% |
| 45-54 | 374 (15%) | 357 (16%) | 13.3% |
| 55-64 | 650 (26%) | 567 (25%) | 12.7% |
| 65+ | 1,160 (46%) | 1,028 (45%) | 18.6% |
| **Ethnic group - n (%)** |  |  |  |
| Asian, Asian British or Asian Welsh | . | 29 (1.3%) | 8.6% |
| Black, Black British, Black Welsh, Caribbean or African | . | 10 (0.5%) | 3.7% |
| Mixed or Multiple ethnic groups | . | 24 (1.1%) | 2% |
| Other ethnic group | . | 4 (0.2%) | 2% |
| White | . | 2,135 (97%) | 83.1% |
| Missing | 2,540 | 71 |  |
| **Highest level of education - n (%)** |  |  |  |
| GCSE / A-level or equivalent | 629 (25%) | 577 (25%) | 48.1% |
| Bachelors degree or higher | 1,801 (71%) | 1,622 (71%) | 33.6% |
| No formal education | 72 (2.8%) | 49 (2.2%) | 18.3% |
| Still in education | 38 (1.5%) | 25 (1.1%) | . |
| **Work activity - n (%)** |  |  |  |
| Retired | 1,143 (45%) | 1,008 (44%) | 21.6%* |
| Full or part-time employment | 1,042 (41%) | 953 (42%) | 47.6%* |
| Self-employment | 128 (5.0%) | 107 (4.7%) | 9.6%* |
| Long-term sick or parental leave (long term sick or disabled) | 45 (1.8%) | 51 (2.2%) | 4.2%* |
| Homemaker (home or family) | 56 (2.2%) | 65 (2.9%) | 4.8%* |
| Unemployed | 18 (0.7%) | 15 (0.7%) | 3.4%* |
| Other | 37 (1.5%) | 29 (1.3%) | 3.1%* |
| Daycare or education | 71 (2.8%) | 45 (2.0%) | . |
| **Smoking - n (%)** |  |  |  |
| No | 2,402 (95%) | 2,148 (95%) | . |
| Yes | 106 (4.2%) | 84 (3.7%) | . |
| E-cigarettes | 32 (1.3%) | 40 (1.8%) | . |
| Missing |  | 1 |  |
| **Allergy - n (%)** |  |  |  |
| No | 1,454 (57%) | 1,297 (57%) | . |
| Yes | 1,086 (43%) | 976 (43%) | . |
| **Asthma medication - n (%)** |  |  |  |
| No | 2,231 (88%) | 1,989 (88%) | . |
| Yes | 309 (12%) | 284 (12%) | . |
| **Diabetes medication - n (%)** |  |  |  |
| No | 2,404 (95%) | 2,151 (95%) | . |
| Yes | 136 (5.4%) | 122 (5.4%) | . |
| **Lung disease medication - n (%)** |  |  |  |
| No | 2,487 (98%) | 2,227 (98%) | . |
| Yes | 53 (2.1%) | 46 (2.0%) | . |
| **Heart disease medication - n (%)** |  |  |  |
| No | 2,315 (91%) | 2,063 (91%) | . |
| Yes | 225 (8.9%) | 210 (9.2%) | . |
| **Kidney disease medication - n (%)** |  |  |  |
| No | 2,528 (100%) | 2,257 (99%) | . |
| Yes | 12 (0.5%) | 16 (0.7%) | . |
| **Immunocompromised medication - n (%)** |  |  |  |
| No | 2,453 (97%) | 2,193 (96%) | . |
| Yes | 87 (3.4%) | 80 (3.5%) | . |
| **Influenza vaccination - n (%)** |  |  |  |
| Yes, I had one | 1,608 (63%) | 1,609 (71%) | . |
| Yes, I'm planning to | 507 (20%) | 245 (11%) | . |
| No | 284 (11%) | 314 (14%) | . |
| I don't know | 141 (5.6%) | 105 (4.6%) | . |
| **COVID-19 vaccination - n (%)** |  |  |  |
| Yes, I had one | 1,287 (51%) | 1,270 (56%) | . |
| Yes, I'm planning to | 370 (15%) | 148 (6.5%) | . |
| No | 637 (25%) | 777 (34%) | . |
| I don't know | 232 (9.2%) | 71 (3.1%) | . |
| *Missing* | 14 | 7 | . |
| **RSV vaccination - n (%)** |  |  |  |
| Yes, I had one | . | 165 (7.4%) | . |
| Yes, I'm planning to | . | 61 (2.7%) | . |
| No, I'm eligible but not planning to | . | 58 (2.6%) | . |
| No, I'm not in one of the eligible groups | . | 1,852 (83%) | . |
| I don't know | . | 93 (4.2%) | . |
| *Missing* |  | 44 |  |

\* Census 2021 England and Wales data as UK data was not available (24). See Text S1 for survey phrasing.

For each indicator, we estimated 95% confidence intervals using bootstrapping (10,000 samples with replacement).

### National surveillance data sources

We extracted published national data on GP ILI consultations, influenza test positivity, and influenza hospital admissions for comparisons with FluSurvey ILI rates among the subset residing in England (17). Data sources were filtered for FluSurvey operational weeks (from w/c 18 November 2024 to w/c 31 March 2025 inclusive) and processed to align data collection dates. As an example, we describe this alignment for w/c 18 November. In FluSurvey this would represent surveys collected between 18 to 24 November, reporting on symptoms experienced in the past week (i.e. ranging from 11 November to 24 November).

#### GP ILI consultations

Weekly ILI consultation rates in GP practices were from the Royal College of General Practitioners (RCGP) GP sentinel surveillance system comprising 2000 GP practices in England. The number of GP consultations for ILI are recorded each week and converted into rates per 100,000 using the number of registered patients among reporting practices as the denominator (20,21). No transformation was required to align data collection; w/c 18 November comprises ILI consultations from 18 November to 24 November.

#### Influenza test positivity

Data on daily influenza PCR positivity as a proportion of all influenza PCR tests conducted in diagnostic laboratories in England was captured by the Second Generation Surveillance System (SGSS). Daily test positivity is estimated using a 7-day rolling positivity rate, estimated using the number of individuals who have tested positive in a seven-day period divided by the total number of individuals who have tested overall in the same period, based on specimen date (20). We therefore extracted data from each Sunday for comparisons with FluSurvey ILI rates. For example, w/c 18 November was represented by 24 November, which comprises tests conducted from 18 November to 24 November.

#### Influenza hospital admissions

Data on weekly influenza hospital admissions (including critical care) were captured by the Severe Acute Respiratory Infection (SARI)-Watch surveillance system. Counts of new test-confirmed influenza admissions are submitted weekly by a sentinel network of NHS trusts in England (20,22) and converted to rates per 100,000 using the trust catchment population of reporting trusts that week (20). No transformation was required to align data collection; w/c 18 November comprises new admissions from 18 November to 24 November.

### Statistical analysis

Analyses were conducted using STATA18 and R version 4.3.2.

#### FluSurvey participation and participant characteristics

We conducted a descriptive analysis of participation and participant characteristics by season. Where comparable variables were available, characteristics derived from UK 2021 Census data were presented alongside (23). Vaccination uptake among the FluSurvey cohort who were living in England and aged 65 and over (using age at 31 March in each season) were compared with national vaccination uptake statistics in the same age-group.

#### Symptom reporting, ILI episodes and illness-related behaviours

We summarised patterns in symptom reporting by season. We then described ILI episodes overall and by season, including symptom profiles, healthcare use, medication, impact on daily life, testing behaviours, as well as differences in health scores.

#### Comparisons with national ILI and influenza surveillance

We calculated FluSurvey ILI indicators for the latest season (2024–25) among the subset residing in England to facilitate cross-correlations with other national surveillance indicators.

We compared the three FluSurvey ILI rates – applying commonly adopted methods to address known biases – in terms of peak week and overall activity. We subsequently examined relationships between each FluSurvey ILI indicator and i) GP ILI consultations, ii) influenza test positivity, iii) influenza hospital admissions (nine comparisons) using Peason’s cross-correlations at time lags of up to +/− 2 weeks.

We computed cross-correlations using the latest season of data. Cross-correlations were implemented using the ccf_boot function in R which uses sieve bootstrapping (1000 simulations) to better account for autocorrelation among the time series.

## Results

### FluSurvey participation

Over two winter seasons, 3057 individuals registered and completed 68,149 symptoms surveys. There were fewer participants in 2024-25 (N=2273 compared with N=2540 in 2023-24), although the season started later and ended earlier. More than half of all participants (N=1757 of 3057) enrolled in both seasons.

Weekly participation is further described in Figure S1. In both seasons, participation peaked in the new year and started to decline after. New registrations were observed in most weeks but comprised a negligible proportion by February. Engagement across the season was relatively high given participants may sign up at any point. Median participation was 18 of 25 weeks in 2023-24 (72% of all weeks) and 15 of 20 weeks in 2024-25 (75% of all weeks).

The median (IQR) time taken to complete a symptom survey was 16 seconds (11,49) in 2023-24 and 32 seconds (20,65) in 2024-25. This varied by symptom reporting: survey duration was 13 seconds (10,19) and 26 seconds (18,38) among those reporting no symptoms for 2023-24 and 2024-25 respectively, and 105 seconds (79,142) and 99 seconds (72,139) where symptoms were reported.

### Participant characteristics

Participant characteristics stratified by season are presented in Table 1 (see Table S1 for the England subset). Characteristics were broadly similar across the two seasons, although we note the large overlap in participation. Most participants lived in England and the cohort differed from the UK general population. For example, there was an overrepresentation of females, White ethnic groups and 45 and over age groups. A larger proportion of participants were additionally retired with higher levels of educational attainment reported.

#### Vaccination

We compared self-reported COVID-19 and influenza vaccination status among the FluSurvey England subset to UKHSA national statistics (25,26). We used the 65 and over cohort – who were eligible for both vaccinations in both seasons – for comparisons. Influenza vaccination uptake was higher among participants at 81.8% in 2023-24 and 89.8% in 2024-25 (compared with 74.9% and 77.8% nationally, respectively). COVID-19 vaccination uptake was also higher among participants at 77.6% in 2023-24 and 84.5% in 2024-25 (compared with 70.4% and 59.3% nationally, respectively). Vaccination status for the full cohort is presented in Table 1.

### Symptom reporting

Of 68,149 surveys, most didn’t report symptoms (77% in both seasons). Symptom reporting peaked in the first week of operation at 30% in both seasons, with a minimum of 17% (towards the end of the season) and maximum of 28% (in week 52) after discarding the first full week of operation.

Rates of individual symptoms, ARI and ILI in each week are presented in Figure S2.

### ILI episodes and illness-related behaviours

ILI episode analyses were conducted in the “active” subset submitting two or more symptom surveys (N_2023-24_=2364; N_2024-25_=2068).

We note caution in direct comparisons of seasonal attack rate given the differences in operational period. Of all ILI reports, 1868 episodes were identified; 1065 in 2023-24 (N_participants_=805, 34.1% of participants) and 803 in 2024-25 (N_participants_=616, 29.8% of participants).

Characteristics of ILI episodes by season, including symptoms, healthcare use, medication, daily routine and testing are presented in Figures 1-2 and Table S2 and described below.

**Figure 1.**
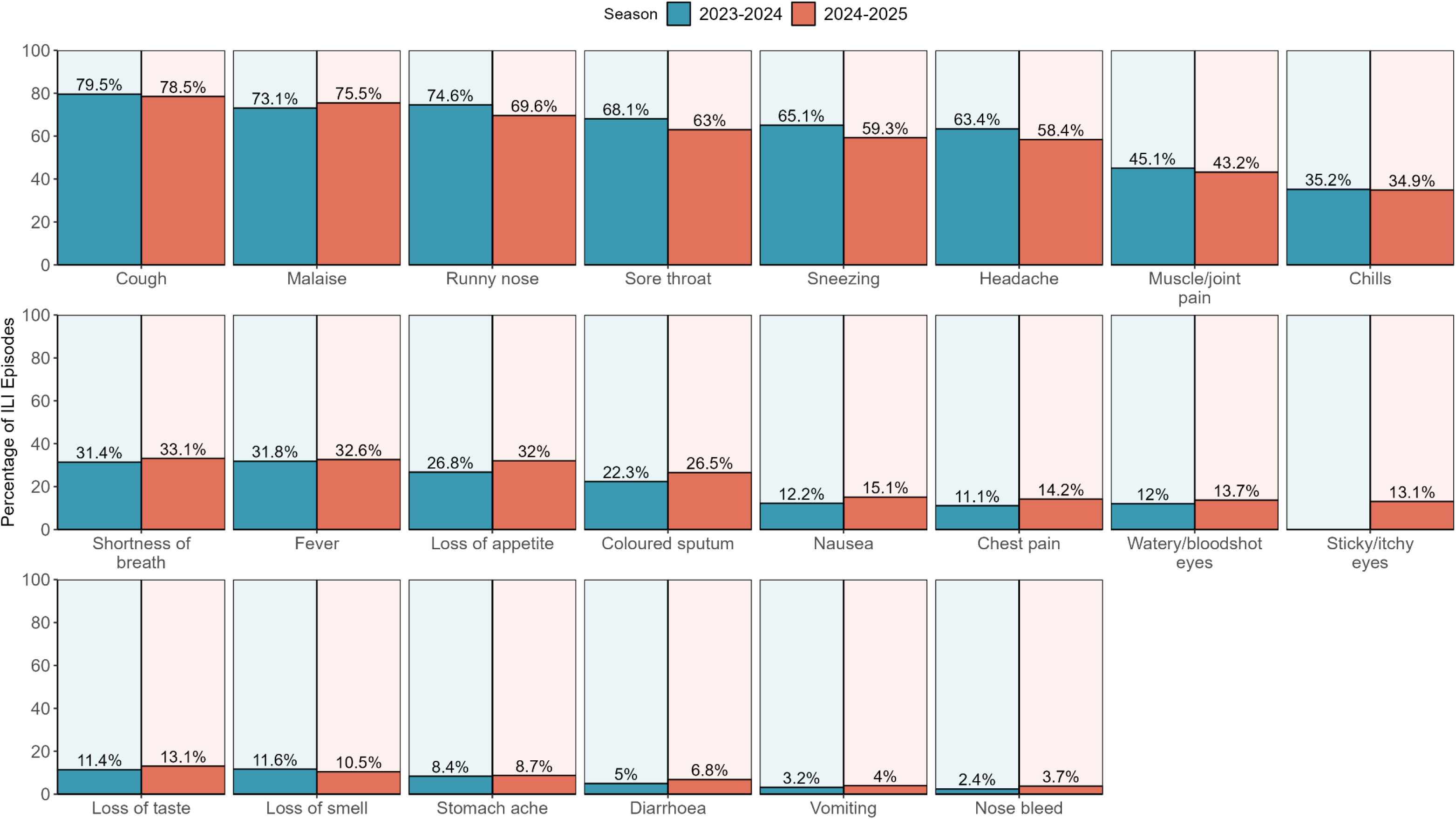
Bar chart displaying the crude proportion of influenza-like illness episodes where the symptom was ever reported (1065 episodes in 2023-24; 803 episodes in 2024-25). *Sticky/itchy eyes was added in the 2024-25 season and therefore data are unavailable for the 2023-24 season*.

**Figure 2.**
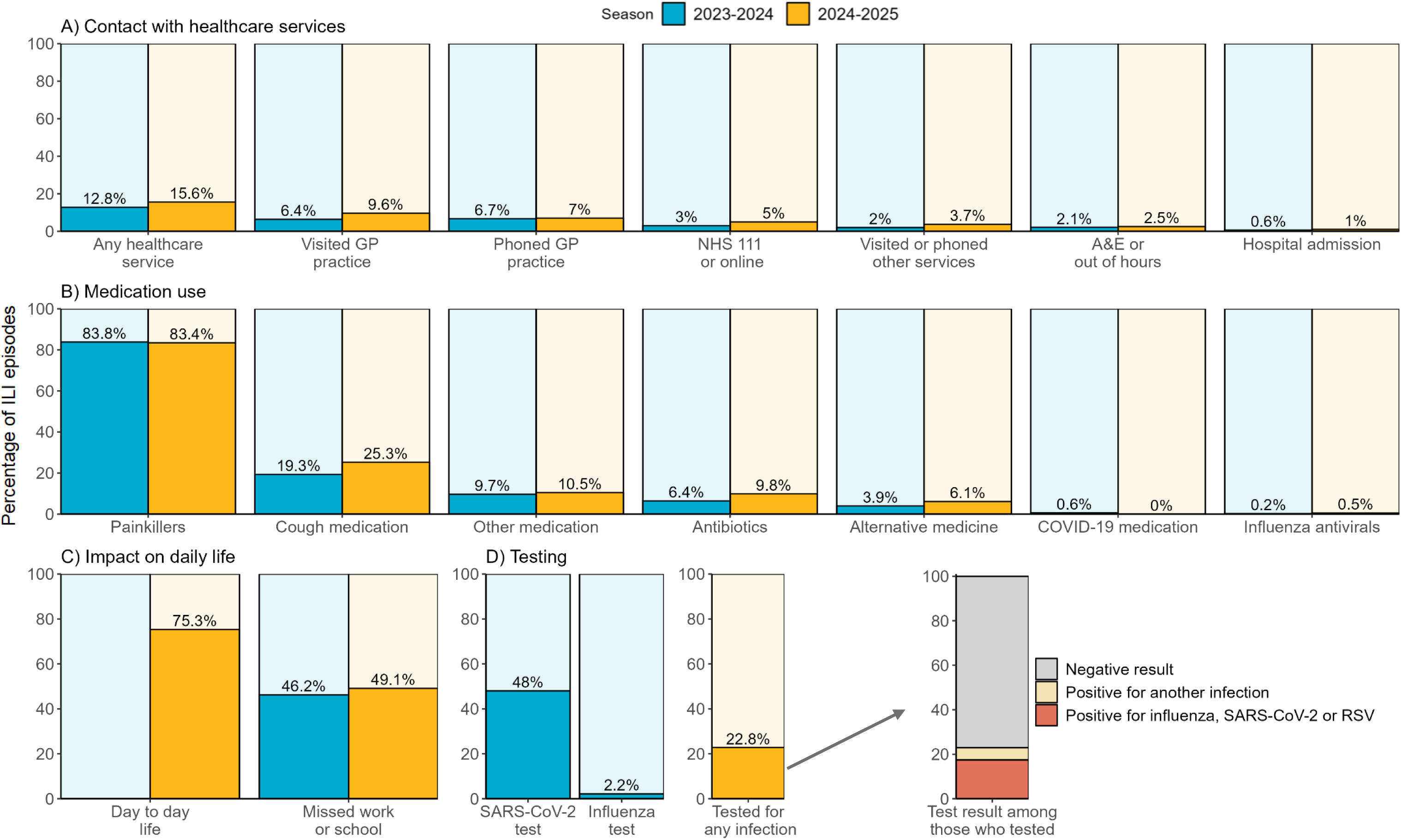
Bar chart displaying the proportion of influenza-like illness episodes reporting healthcare service use, medication use, impact on daily life, and testing (1065 episodes in 2023-24; 803 episodes in 2024-25). *Testing was reported as use of a COVID-19 or influenza test in the 2023-24 season, with no results indicated. This was rephrased in 2024-25 to include any test and test result. No suitable measure was available for impact on daily life in the 2023-24 season*.

#### Symptoms

Symptom profiles were broadly similar across both seasons; the top symptoms (reported in >50% episodes) were cough, malaise, runny nose, sore throat, sneezing and headache. Non-respiratory/systemic symptoms were not uncommon: 21% of ILI episodes reported gastrointestinal symptoms (2023-24: 20%; 2024-25: 23%); 23% reported ocular symptoms (2024–25), and 15% reported loss of taste or smell (2023-24: 15%; 2024-25: 15%).

#### Healthcare contact

Most episodes did not result in contact with healthcare services; 14% reported contact with any healthcare service (2023-24: 13%; 2024-25: 16%), of which the most common was a GP visit (2023-24: 6.4%; 2024-25: 9.6%). Among those visiting a GP, the median (IQR) delay from symptom onset was 7 days (4,12) in 2023-24 and 5 days (3,9.5) in 2024-25. Less than 5% used NHS 111 or online services, visited A&E or used out of hours services; less than 1% were admitted.

#### Daily life

The majority of ILI episodes reported impact on daily life (75.3%; 2024-25 only). Among those in employment or education, 47.4% missed work or school (2023-24: 46.2%; 2024-24: 49.1%). Overall, there was a median relative decrease of 29% compared with health scores when the participant reported no symptoms. The median health score was 90 when the participant reported no symptoms (2023-24: 90; 2024-25: 89) and the median minimum health score across all ILI episodes was 60 (2023-24: 60; 2024-25: 59).

#### Testing

Although questions were phrased differently by season, changes in testing behaviour were still apparent. Approximately half (48%) of all ILI episodes reported testing for SARS-CoV-2 in 2023-24, of which 97% used LFDs. Whereas in 2024-25 less than a quarter (23%) reported taking any test (including SARS-CoV-2).

Of the 183 episodes reporting taking a test in 2024-25, 42 (23%) were positive for any infection, including 32 positive for ≥1 of influenza, SARS-CoV-2, RSV (N=14 influenza only; N=15 SARS-CoV-2 only). No test result data were available for the 2023-24 season.

#### Medication

The majority of episodes used medications (2023-24: 89%; 2024-25: 90%). Most reported use of painkillers (2023-24: 84%; 2024-25: 83%), followed by 22% reporting use of cough medication (2023-24; 19%; 2024-25: 25%). Antibiotics and alternative medicine were each taken in <10% of episodes. A negligible proportion reported receiving influenza antivirals or COVID-19 therapeutics.

### Comparisons with national ILI and influenza surveillance

#### FluSurvey ILI rates

FluSurvey ILI rates are presented in Figure 3, with comparator data sources plotted alongside in Figure S3. Overall activity was broadly similar with the same peak week across the three ILI rates (Figure 3). Peak ILI rate varied in magnitude across the three indicators: crude ILI rates peaked at 73.3 per 1000 (95%CI 58.7-88.1), with a higher peak activity observed for weighted rates at 99.5 per 1000 (95%CI 74.8-126) and a lower peak activity for crude ILI rates at 53.2 per 1000 (95%CI 40.7-66.5). Weighted ILI rates were higher across most weeks although confidence intervals were wide. ILI rates following QC were lower in particular during peak activity.

**Figure 3.**
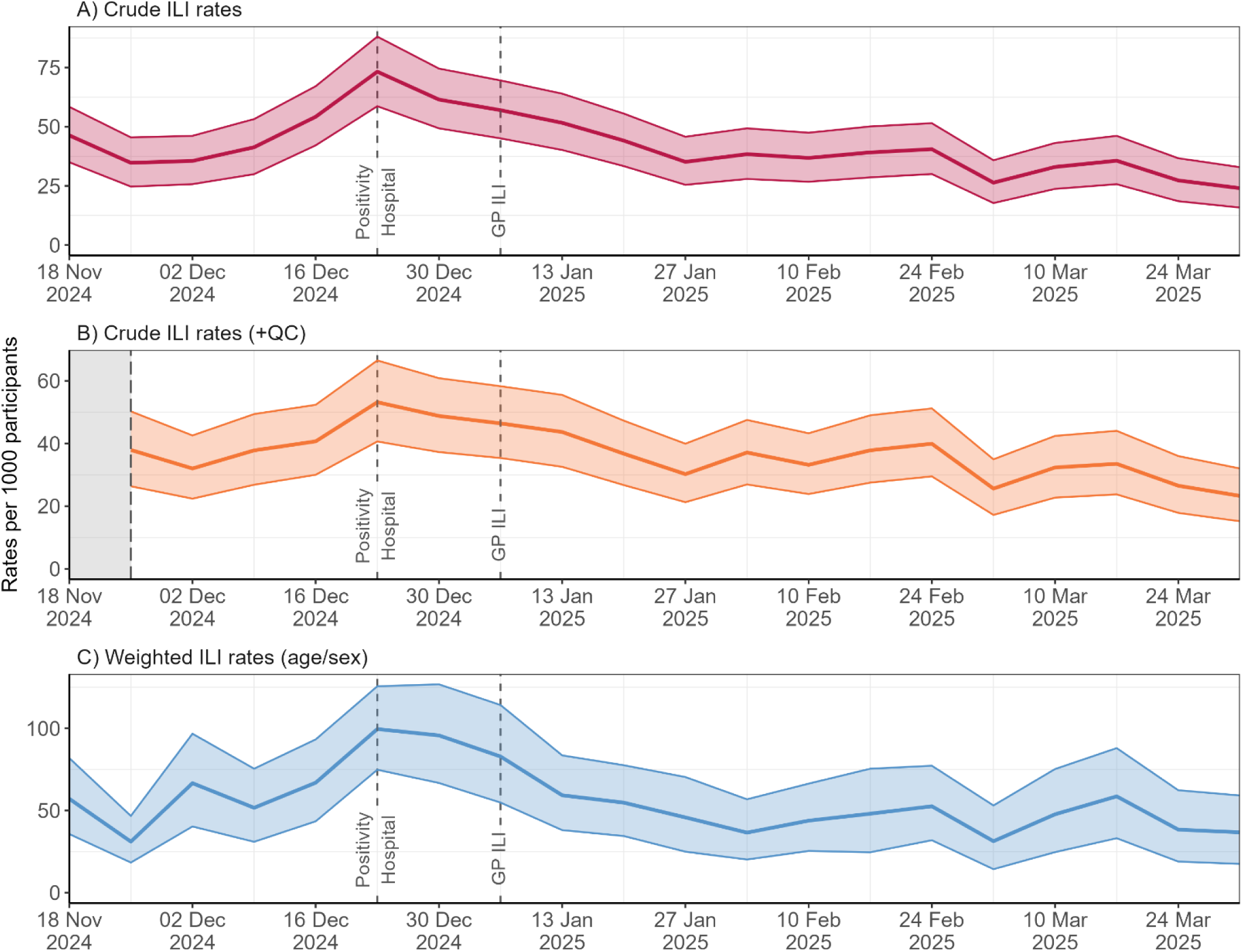
Comparison of the three FluSurvey influenza-like illness (ILI) rates in the 2024-25 season (England). FluSurvey indicators include a) crude rates, b) crude rates with the first survey omitted, and c) age/sex standardised rates. Ribbons display bootstrapped 95%CI. Data are plotted by week commencing date according the date the survey was submitted. Peak weeks reported in the surveillance of influenza test positivity, influenza hospital admissions and GP ILI consultation rates are annotated with a dotted line (3)*. Daily test positivity peaked 30 December (representing collected 7 days prior) and is annotated as w/c 23 December for consistency with the other weekly data*.

Peak FluSurvey ILI activity coincided with peak influenza hospital admissions and influenza test positivity and preceded GP ILI consultation rates by two weeks.

#### Cross-correlations with national surveillance

Correlations of FluSurvey ILI rates with GP ILI consultations, influenza hospital admissions and test positivity are presented in Figure 4.

**Figure 4.**
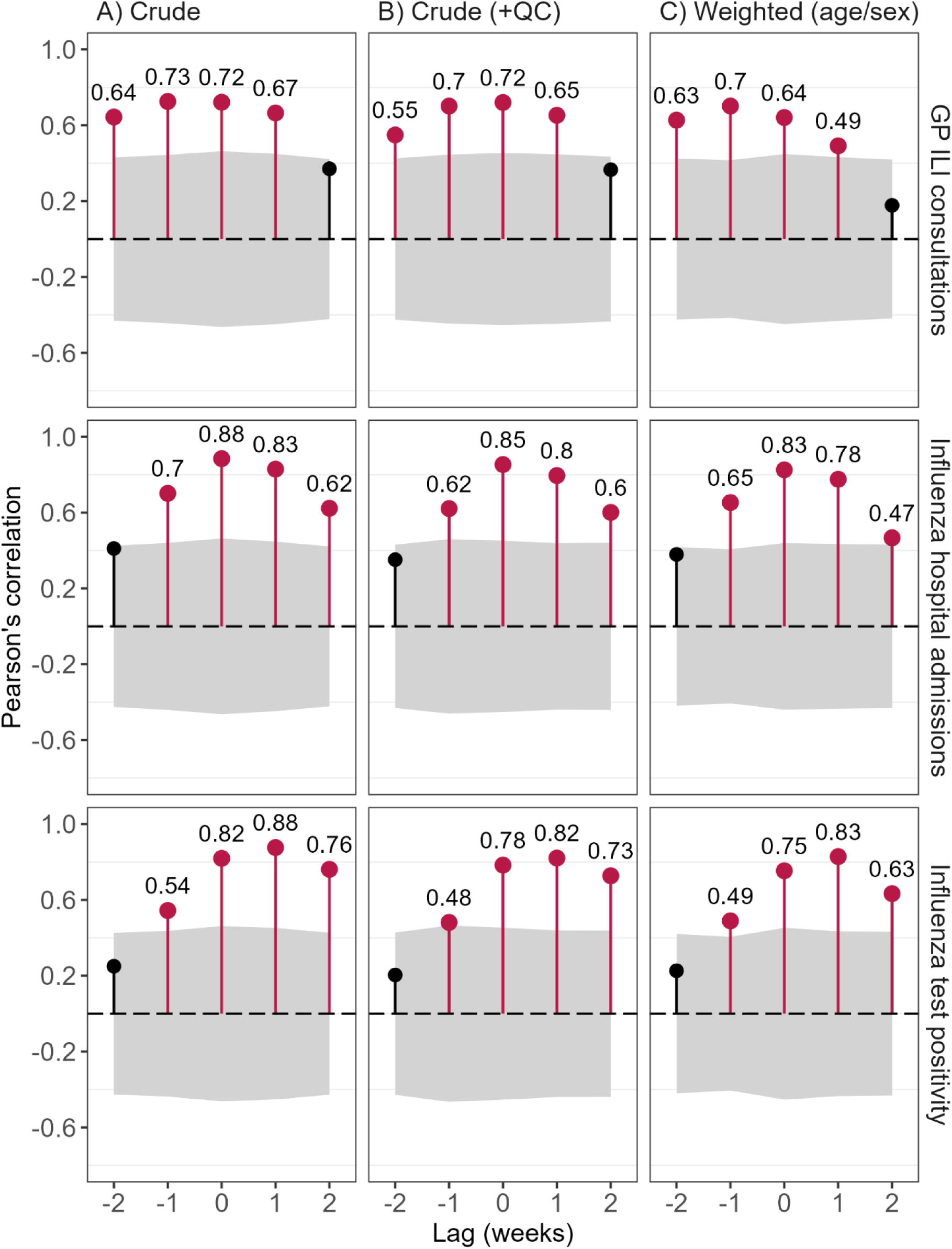
Cross-correlations of FluSurvey influenza-like illness (ILI) rates (England) with influenza test positivity, GP ILI consultations, and influenza hospital admissions in winter 2024-25. Pearson’s correlations were calculated at time lags of +/− 2, +/− 1 and 0 weeks. *95% confidence limits under the null were calculated using sieve bootstrapping and are shaded in grey. Statistically significant correlations are displayed in red with correlation coefficients annotated. Time lags represent FluSurvey ILI rates shifted according to the value represented, i.e. –2 weeks would represent rates 2-weeks prior*.

Strong and significant correlations (r>0.7) of FluSurvey ILI with national surveillance indicators were observed across all analyses and strongest for crude ILI rates. For GP ILI consultations, these were observed at –1 week and 0-weeks and were slightly larger at –1-week (r=0.73), suggesting a leading relationship of FluSurvey ILI with GP ILI consultations.

Correlation coefficients were stronger in analyses of both influenza hospital admissions and test positivity and suggested similar timing with hospital admissions and lagging relationships with test positivity. For hospital admissions, correlations were largest at the same time point (r=0.88), with strong correlations additionally reported 1-week either side. For test positivity, correlations were largest at 1-week (r=0.88) with strong correlations again observed 1-week either side.

## Discussion

Using the UK’s longest-running participatory surveillance tool – FluSurvey – we characterised ILI and illness-related behaviours following the COVID-19 pandemic over two winter seasons. Although the large majority of ILI did not report contact with healthcare services, we saw wider impacts on self-reported health and daily routine including absenteeism. Cross-correlations indicated strong correspondence of ILI with national influenza surveillance (GP consultations, hospital admissions and test positivity), despite inherent differences among the populations represented in each system.

### High ILI prevalence, low healthcare-seeking behavior, and broader community burden

A high ILI attack rate and low proportion of medically-attended illness aligns with findings from the InfluenzaNet consortium, analysing similar surveys deployed across Europe in both pre-pandemic and pandemic periods (7,27). Interestingly, these analyses found that UK (FluSurvey), Switzerland and the Netherlands report lower healthcare-seeking behaviours than other countries in Europe, possibly explained by differences in public health attitudes, healthcare access, and workplace policies. Since risk groups for severe disease comprise the very young and old in many cases, their under-representation typically observed among participatory surveillance may also impact our estimates. Nevertheless, age/sex standardisation resulted in similar conclusions (27), and a low percentage of respiratory tract infections consulting with healthcare settings has been reported using a representative sample in the UK (28). Healthcare-seeking behaviour may also be underestimated if severe presentations are less likely to participate in FluSurvey. However, as the majority of ILI is self-limiting, with guidelines directed towards staying at home where possible (29), we would not expect them to change substantially.

Irrespective of healthcare use, we saw notable impacts on daily life due to an ILI episode, with a 29% decrease in self-reported health scores, 75% reporting changes in daily routine, and 47% missing work or school. The most prevalent symptoms were mostly mild; however, the majority reported taking medication, and fever and lower respiratory symptoms were not uncommon, nor were GI or ocular symptoms. As most surveillance data lacks granularity on daily life impacts and symptom profiles, these findings provide valuable information on respiratory illness circulating in the community for public health response. For example, monitoring ocular symptoms in the context of conjunctivitis and avian influenza (30).

To our knowledge, testing behaviour has not been examined among the general population in recent seasons. Here, we saw a drop in testing in 2024-25 in comparison to 2023-24, where roughly half of all ILI reported testing for COVID-19 due to symptoms (of which almost all were LFD). Although there were changes to national policy on 1 April 2024 (31), these were applicable to high-risk settings, and changes to community testing in the general population preceded both seasons. With LFD innovations for other respiratory viruses (32), as well as increased acceptability and literacy of home testing, it will be interesting to monitor patterns in community testing for influenza and RSV in coming seasons.

### Strong correlations with national surveillance

Since the ECDC ILI definition is broad and can capture COVID-19 and other respiratory illness (15), it was reassuring to see strong correlations and similar timing of peak epidemic activity with national influenza surveillance. Our findings are in line with numerous other studies comparing participatory ILI indicators with national surveillance, where moderate or strong correlations were reported (4,7). However, these studies used data collected prior to COVID-19, and this is the first to compare with hospital admissions representing more severe disease.

Given FluSurvey ILI indicators comprise community symptoms, we would expect these signals to precede or correspond with indicators requiring healthcare service use. We saw strong correlations at the same time point and one week either side in most cases, except for test-positivity where these were unexpectedly slightly later. Maximum correlations led GP ILI consultations (r=0.73), coincided with hospital admissions (r=0.88), and lagged test positivity (r=0.88). Nevertheless, peak activity either preceded (for GP ILI surveillance) or coincided (for test positivity and hospital admissions) with other systems.

Comparing timeliness is complicated through FluSurvey comprising surveys reporting on symptoms in the previous week – hence including a wider time range. The comparator data sources are also reported according to initial presentation (e.g. specimen date for test positivity) and retrospectively updated through the season as data becomes available. Here, we extracted end-of-season data and compared with FluSurvey metrics according to the date the survey was submitted. If FluSurvey was analysed by onset date, or surveys were analysed more frequently, we may observe earlier signals for FluSurvey than reported here.

Surveillance systems are also characterised by different designs and populations, capturing patterns across the spectrum of disease to inform public health action, each with different timings following symptom onset. Younger age groups are comparably over-represented in primary care, whereas the elderly are over-represented in hospital systems (3). Seasonal activity can vary by age group; for example, in 2024-25 influenza B activity was observed later in the season among those aged under 45, with minimal signal in older age groups (3).

Nevertheless, where populations may overlap, systems also differ in design. Test positivity is based on PCR tests conducted nationally in several settings, whereas hospital admissions comprise test-confirmed admissions confirmed through a wider range of diagnostic tests and reported by a network of NHS trusts. Circulating pathogens can also impact test positivity and ILI rates (33). Finally, winter pressures and bank holidays may affect the overall timing from these systems. We saw this in the 2024-25 season where GP ILI consultations dipped w/c 23 December, coinciding with peak influenza activity elsewhere (3). These characteristics may explain some of the unexpected patterns in timing of activity across the timeseries, although overall they aligned well across similar time points.

### ILI methodology

Correlations for crude rates were strongest in all cross-correlation analyses, and weighted rates were weakest in many cases – particularly with hospital admissions. This may reflect closer alignment among populations represented for crude rates, and we would not conclude that weighted metrics are inferior for influenza surveillance. Instead, we recommend that surveillance indicators are interpreted in the context of the populations and season under study. Hence, the strength and timing of relationships may vary by season based on epidemic characteristics, population cohort, circulating viruses, and healthcare pressures.

While weighted rates were highest, the QC method (discarding all first reports across the season) reduced activity during peak periods. This approach may be less impacted by reporting bias but could reduce overall signal if participants are then less likely to report ILI in following weeks. Given symptom reporting peaked in the first week of operation, where atypical patterns in ILI were also notable, we would instead recommend weeks with a high number of new signups are interpreted with caution.

Although crude rates performed well overall for near-real time surveillance, increasing the diversity among the cohort across under-represented groups is critical. More sophisticated weighting methods may additionally be explored to account for the characteristics of FluSurvey with respect to the general population, such as vaccination status. By characterising the FluSurvey cohort, we are better equipped to interpret emerging trends in the context of other influenza surveillance, as well as supporting situational awareness where other systems are under pressure.

### Strengths and limitations

A key strength of FluSurvey is the long-running, flexible, and scalable format, where content and frequency can easily be amended in response to public health need. Through the application of a standardised and validated symptom-based definition, it can support international surveillance and may be more robust to changes in policy or where other systems are under pressure. The survey additionally offers insights into health-seeking and illness-related behaviours, providing context for other systems during winter season.

There are several limitations to note. First, as an online voluntary platform, the cohort have a distinct demographic and were not representative of the UK population. We applied age/sex post-stratification weights but due to low numbers these required large categories for age-groups (e.g. <45years) and we were unable to account for other factors such as ethnicity. Second, as a symptom-based measure, ILI is practical and low-cost but is not specific to influenza. We are careful to communicate this appropriately in outputs, but the integration of self-testing could allow additional insights on test-confirmed illness. Third, reporting bias can be introduced through individual self-selection, as well as recall bias through self-report. We implemented quality control methods to mitigate both, but it is likely that residual bias remains. Fourth, cross-correlations are typically more stable over longer time-series. Future work should attempt to repeat these using a wider range of other data sources with longer operating periods. In addition, we note that the 24-25 season analysed was characterised by high influenza activity and comparably low COVID-19 activity. If there were multiple respiratory illnesses with compatible presentations in high circulation, correlations may be weaker due to the lower specificity of ILI (33).

## Conclusion

The majority of influenza-like illnesses reported to FluSurvey do not contact healthcare due to symptoms but experience wider impacts on daily life. FluSurvey ILI corresponded well with other national influenza surveillance tools and provided broader context on community illness and health-seeking behaviours, supplementing the surveillance of influenza and other respiratory viruses for public health response. The continued monitoring of ILI characteristics will be important for the timely detection of unusual trends, and the integration of virological testing may strengthen this further. Expanding and broadening participation among the cohort remains a priority to enable more representative insights.

## Data Availability

Summary data used for cross-correlation analysis are publicly available https://www.gov.uk/government/statistics/national-flu-and-covid-19-surveillance-reports-2024-to-2025-season. Authors cannot make the underlying dataset publicly available for ethical and legal reasons.

## Acknowledgements

We are grateful to members of the Respiratory Virus Section Surveillance team, in particular Chinelo Obi, Alex Allen, Jamie Lopez-Bernal, Kiara Assaraf, Freja Kirsebom, Catherine Quinot and Suzanne Elgohari who produce data sources included in UKHSA’s weekly influenza and COVID-19 surveillance report used for this analysis.

## Ethics

Data are processed under the UK General Data Protection Regulation (UK GDPR) lawful bases that relate to public interest and public health: Article 6(1)(e) and Article 9(2)(i). Separately, common law consent is obtained to meet the legal and ethical requirements for processing confidential patient information, in line with the common law duty of confidentiality.

## Conflict of interest

The Immunisations and Vaccine Preventable Diseases division at UKHSA has undertaken post-marketing surveillance and regulatory analyses requested by vaccine manufacturers for which cost-recovery charges have been made. No other conflicts of interest have been declared.

## Funding statement

This work was funded by the UK Health Security Agency. No external funding was received.

## Supporting information

Table S1. FluSurvey participant characteristics in the 2023-24 and 2024-25 seasons (England subset only).

Table S2. Characteristics of influenza-like illness episodes by symptoms, healthcare use, medication, testing, and impact on daily life among FluSurvey participants.

Figure S1. Bar chart displaying the distribution of participants and new FluSurvey participants by week in each season.

Figure S2. Rates per 1000 FluSurvey participants reporting any symptom, individual symptoms, and meeting the influenza-like illness (ILI) and acute respiratory infection (ARI) case-definition in the 2023-24 and 2024-25 season.

Figure S3. FluSurvey influenza-like illness (ILI) rates (crude, crude with first survey omitted, age/sex weighted rates) plotted alongside surveillance of B) GP ILI consultations, C) influenza hospital admissions, D) influenza test positivity.

Text S1. Supplementary Text.

